# Probiotics and synbiotics administered to young infants: perceptions and acceptability amongst carers and healthcare workers in Western Kenya

**DOI:** 10.1101/2024.12.20.24319230

**Authors:** Mary Iwaret Otiti, Florence Achienge, Sevim Zaim, Helen Nabwera, Simon Kariuki, Stephen Allen

**Affiliations:** Kenya Medical Research Institute (KEMRI) Centre for Global Health Research, Kisumu, Kenya; MSc Humanitarian Studies; Liverpool School of Tropical Medicine, Liverpool; Department of Clinical Sciences, Liverpool School of Tropical Medicine, Pembroke Place, Liverpool

**Author notes:** Corresponding author: Mary Iwaret Otiti. Authors emails: FA, SZ, HN, SK, SA.

**Keywords:** Malnutrition, probiotics, synbiotics, acceptability, barriers, perceptions, carers, Peer Mothers

## Abstract

**Background:** A contributory factor to childhood undernutrition is poor gut health occurring within the first 6-12 weeks of life despite exclusive breast feeding. Pro/synbiotic administration may protect gut health. A qualitative study was conducted amongst mothers/carers and healthcare workers (HCWs) to explore their perceptions and the acceptability of pro/synbiotics administration in early life.

**Methods:** This study was nested within a randomised, open, clinical trial of pro/synbiotics with 32 doses administered under supervision to infants between age 0-5 months in western Kenya. Semi-structured interviews were conducted with mothers/carers, Peer Mothers and health care workers (HCWs) selected by purposive critical and key informant sampling. Interviews were transcribed and analysed using a thematic coding framework.

**Findings:** Satisfaction with pro/synbiotic administration was very high amongst all three groups. Commonly perceived benefits included protection from diseases, healthy growth of the infant and improved appetite. The main barriers were working mothers and other commitments making it difficult to stick to scheduled administration visits, adverse judgment and opinions in the community and lack of engagement of fathers. Insights were gained into different means of administering pro/synbiotics to young infants. Triangulation of findings of the mothers/carers with HCWs showed that most identified motivations and challenges were similar.

**Conclusions:** Pro/synbiotic administration was well-accepted by mothers/carers and HCWs and generally perceived to have health benefits. Administration of pro/synbiotics by mothers/carers themselves to their infants may be feasible and overcome logistical challenges. Greater efforts to sensitise and engage fathers and communities would likely be critical for a community-based program.

## Introduction

Childhood undernutrition remains a global concern with wasting estimated to affect 6.8% and stunting 22.3% of all children under 5 years of age in 2022. Ninety-seven percent of all wasted and 95% of all stunted children live in Asia or Africa [1]. Several nutrition-specific interventions including promoting exclusive breastfeeding, nutritional supplements, immunomodulators, and improved water, sanitation and hygiene (WASH) have had limited impact on preventing malnutrition [2–4]. This has renewed interest in the abnormal structure and function of the gut termed “environmental enteric dysfunction” (EED). EED a sub-clinical condition characterized by damage to the small intestine thought to be due to colonisation of the gut by a range of enteropathogens as a result of poor sanitation and hygiene. [5–7]. EED likely impairs growth due to impaired nutrient digestion and absorption and increased systemic inflammation. EED can occur within the first 6-12 weeks even amongst exclusively breastfed infants [8,9].

Probiotics are live non-pathogenic microorganisms, which, when administered in adequate amounts, confer a health benefit on the host [10]. Probiotics may provide colonisation resistance against enteropathogens and also improve gut health through several mechanisms [11, 12]. Prebiotics support the growth of beneficial microorganisms conferring a health benefit to the host [13]. Synbiotics combine probiotics with prebiotics [14].

The administration of pro- and synbiotics to young infants in poorer communities may be feasible, safe and bring health benefits [15–17]. However, the perceptions regarding nutritional supplements and their acceptability amongst parents/carers and healthcare workers (HCWs), especially in exclusively breast-fed infants, needs to be considered in the development of this intervention for community settings where undernutrition is common. We assessed the perceptions regarding pro/synbiotic administration and their acceptability amongst parents/carers, Peer mothers and HCWs in a low-resource setting in western Kenya.

## Methods

### Study setting and population

This qualitative study was nested within the PRObiotics and SYNbiotics in infants in Kenya (PROSYNK) randomized, open trial in Homabay county, Nyanza Province, western Kenya [Trial registration: PACTR202003893276712; 18]. The main aims of the study were to determine whether administration of pro/synbiotics in newborns during the first 0-5 months of life would improve gut health and growth. Stunting occurs in 13% of under-fives in this region [19]. Residents are predominantly of the Luo ethnic group, whose common livelihoods include agriculture, fishing, and microenterprises.

Between October 2021 and January 2022, newborns delivered at the Homabay County Teaching and Referral Hospital with birthweight ≥2000g, who had taken at least one feed well and in whom there were no health concerns were recruited within the first 3 days of life. They were randomized to one of three groups (either a probiotic or one of two synbiotics) or a control arm (no intervention). Pro/synbiotics were presented as powder in capsules and were given once daily for the first 10 days and then weekly to age 6 months (a total of 32 visits/child). Each capsule was transported to the infant’s home in a cold box. After hand sanitising and using appropriate hygienic measures, the contents of the capsule could be sprinkled directly into the infant’s open mouth before feeding or mixed in a clean container with sterile water. All administration was supervised by a member of the research team. Mothers/carers also received support from trained Peer Mothers who were selected from the community having participated in a previous study involving low birth weight babies [20]. Both research staff and Peer Mothers advised mothers/carers on newborn and infant care practices during visits for pro/synbiotic administration. Community sensitization was conducted in the communities prior to recruitment and enrollment into the study.

Compliance with follow-up visits and pro/synbiotic administration was high with 85% home visits completed and 83% doses administered successfully. Follow-up visits occurred mainly in infants’ homes with some administration occurring during routine hospital follow-up (e.g. for immunisation). Consistent with the Kenya national guidelines for research [21], face-to-face visits for pro/synbiotic administration under supervision were maintained with the required precautions during the COVID-19 pandemic; however, follow-up was done by mobile phone calls in the control arm during this period. Monitoring for serious adverse events was undertaken throughout the study and none were attributed to pro/synbiotic administration.

### Sampling procedure

The study was undertaken mainly in the rural community in Nyanza region. Three participant subgroups were purposefully identified: 1) mothers/carers (mothers or carers of infants receiving or who had received a pro/synbiotic in PROSYNK and remained engaged in the trial); 2) HCWs (nurses and doctors from the county hospital and study clinicians); and 3) Peer Mothers. Recruiting mothers/carers of infants enrolled in PROSYNK captured diverse perspectives and experiences at various stages of the study, from recruitment to completion of administration at age 6 months. HCWs were included to explore how pro/synbiotic administration could be implemented alongside recommended infant feeding practices including exclusive breast feeding. Peer Mothers provided a perspective from the community. Mothers/carers, HCWs, and Peer Mothers who declined to give consent, were ill, or mothers/carers’ whose children were ill at the time of interviews were excluded.

### Data collection

Following an explanation of the study objectives and procedures and securing informed consent, semi-structured, in-depth interviews (IDIs) were undertaken during July 2021. IDIs with mothers/carers and Peer Mothers took place in the community at a convenient time which often coincided with home visits for pro/synbiotic administration. IDIs were done in the language which the participants felt the most comfortable expressing themselves (Kiswahili, Dholuo or English). IDIs with HCWs at the hospital were conducted at a convenient time and mostly remotely via WhatsApp mobile application as this was during the COVID-19 pandemic period. A stable internet connection was established at the hospital and IDIs conducted in English. Topics addressed during IDIs included the acceptability of pro/synbiotics and administration processes, perceived benefits and barriers to the intervention, knowledge of infant feeding practices and infant care practices.

### Data management and analysis

IDIs were recorded and stored on passcode-locked digital voice recorders and supplemented by field notes. The interviews were transcribed and translated into English by a research assistant who was a native speaker of both Kiswahili and Dholuo. All transcripts were reviewed by two study investigators (FA, SZ), for clarity and translation quality allowing the investigators to inductively identify patterns prior to analytic coding.

The Health Belief Model (HBM) [22,23] was chosen to achieve a better understanding of an individual’s approach to a health-related decision. Data analysis adopted the ‘thematic analyses method’ approach for the systematic analysis of key and recurring themes. Although time constraints prevented triangulation using different methods of data collection, patterns of convergence between the study groups were investigated during analysis.

Audio recordings were transcribed using Expresscribe software into a MS Word transcription template. Themes were generated first from the interview guide and later codes were developed from responses. The iterative analysis and arrangement of the data into identified themes (and consequent codes) were achieved by re-reading and familiarisation of the data by the researcher and was facilitated by the NVivo software. A code sheet was developed from the first few source documents and later a master code sheet was developed. SZ and a Research Assistant independently coded the responses and thematically analysed the data to draw both descriptive and explanatory conclusions about participants perceptions on perceived benefits or barriers of the intervention and infant care practices. The study team familiarized themselves with the data by re-reading the transcripts to identify recurring issues, inconsistencies, and possible categories; creating a coding framework and then coding using a mixture of deductive codes from our topic guide and inductive codes emerging from the data which the study then applied to all transcripts. The codes were then searched to identify themes, which were reviewed and defined. The coding process was an iterative process involving changing and adding to them as more data became available. Participants would be recruited to each of the three sample groups until it was considered that data saturation had been achieved where no new information would emerge from additional IDIs.

All consent forms were kept in a secure locker. All recordings and transcriptions were kept on lockable devices which could only be accessed by key research staff. All audio recordings were deleted as soon as the interviews were transcribed and no participant identifiable information was written on the transcription sheets. No data were shared outside the research team.

### Ethical statement

Ethical approval was obtained from the Kenya Medical Research Institute’s Scientific and Ethics Review Unit (KEMRI/SERU/CGHR/320/3917) and the Liverpool School of Tropical Medicine Research Ethics Committee (19-048). Signed/thumbprinted informed consent was obtained from all participants prior to data collection. Due to high levels of illiteracy in the study communities, participants incapable of providing written informed consent were verbally guided through the study consent procedures and indicated their consent with a thumbprint.

## Results

Thirty-three participants were interviewed across the three sample groups from different religions and geographical areas within Homabay County. This number achieved data saturation.

Amongst 14 mothers/carers, ten were aged 20-34 years and four 35-50 years; six had not completed primary education, five had incomplete or had completed primary education and three had completed secondary education; all were married except for four who were either single, separated or widowed. Three mothers/carers were housewives/unemployed, eight ran small scale businesses, and three were employed.

Amongst the 12 Peer Mothers, seven were aged 20-34 years and five 35-50 years; three had primary education, seven had incomplete high school, two had completed high school or had higher-level education; all but one were currently married.

Seven key informants included 5 study clinicians and two 2 HCWs aged between 25 and 60 years.

### Process of pro/synbiotic administration

During the initial few days following recruitment into the trial, pro/synbiotic administration to newborns was done almost solely by trained Peer Mothers and clinical officers. However, they would slowly pass on the responsibility to the mothers/carers by teaching them the safe and appropriate technique and eventually letting the mothers administer the pro/synbiotics under supervision. All three participant groups found the administration process of the pro/synbiotic relatively easy. Hygiene awareness during the administration among Peer Mothers was very high and the process was highly standardised as described by one Peer Mother:

> *“When you get to the house of a mother, you have to sanitize your hands, the cooler box carrying the supplement, after that, you clean it using a paper towel, after that you take another paper towel and cover the top of the cooler box because that is what you would use as the surface, then you put on your gloves, that is when you open the cooler, after opening it, you take the supplement and put it on the surface that you had set, then you wipe the cup that you are going to use, after that, you open the supplement and put it in the cup then if you are supposed to use sterile water you use, or the mother might express the breastmilk, but before that, she has to sanitize her hands and wash her hands on a running water, so that we can prevent germs from getting to the child” (Peer Mother 006).*

### Administration of pro/synbiotics by mothers

There were mixed views regarding whether the HCWs and Peer Mothers believed that the mothers/carers could administer the pro/synbiotics themselves. While most of the HCWs were positive that the process was easy enough for mothers/carers to administer the pro/synbiotics on their own, Peer Mothers were more cautious. Nine (75%) Peer Mothers expressed concerns in terms of maintaining good hygiene standards and compliance in the absence of the trial staff. This conflict is demonstrated by the following two quotes:

> *“They can be administered by the mother themselves if the mother is enlightened and is taught and you have observe and you have done the demonstration in her presence; you do it, and then you allow her to do it in your presence, then you are sure that this mother will do it after she understands the importance of participating.” (MOH 001)*

> *“It is not easy because you don’t know what might happen when you leave for them the supplement, you cannot be sure whether the mother has given it or not. And maybe when you left it for them, they would want to taste how it is, at times her other child can unwrap it, so when you left it, we are not certain if they will give it because even with the medicines they are given you will find that there are some mothers that don’t give them to their children, that is why we go there and ensure that the child gets all of it and take everything back to the facility” (Peer Mother 002)*

### Determining factors for method of administration

Opinions on the preferred method of administration of the pro/synbiotics varied among all participants. Most Peer Mothers and HCWs reported that their method of choice would vary on a case-by-case basis and were hesitant to settle on one specific method. However, most agreed that pouring the powder directly into the infants’ mouth was better tolerated with older infants as described in the following two quotes:

> *“Personally, I don’t have a preferred method that I will use. But I will just say at an early stage between the first 10 days, it’s always hard to administer the supplement directly to the mouth. In that event, it actually chokes the participants. that is where we prefer using sterile water and expressed breast milk during the first 10 days of life on up to three weeks of life. So, it’s okay at that point.” (HCW 005)*

> *“So, in terms of delivering them, I use the baby’s age. Because when they are still young, maybe at enrolment on day one or day two, delivering it as powder at times becomes tricky. So, it’s easier when you mix it with sterile water, but then at around the age of between four and five to six weeks, that’s when we can now start giving it directly to the baby’s mouth. So personally, I use both but in terms of the age of the baby.” (HCW 002)*

Both HCWs and Peer Mothers found direct administration as the most convenient and reliable method in older infants:

> *“The method that we have at the moment is the best, administering it in the mouth directly, because that will convince us for sure that the infant has taken.” (MOH 001)*

Six (42%) mothers/carers agreed that direct administration was a better confirmation that the complete dose was taken by their baby as evidenced in the following two quotes:

> *“I am choosing [direct administration] because, when placed in the bottle, you may find that there are some remains in the bottle” (Mother 001, IDI)*

> *“… when it is not mixed, there are some content of the medicine that remains on the bottle” (Mother 010)*

In contrast, some mothers/carers mentioned that their babies sometimes could struggle to consume the pro/synbiotic when given directly in powder form.

> *“With the one given directly, at times she finds it hard to consume it.” (Mother 007)*

The importance of involving the mother/carer in decision-making regarding administration was highlighted by one HCW:

> *“The mother sometimes would suggest, and whenever they suggest, we go with what they suggested. Sometimes you go to administer the supplement and the mother would ask you to just give them the cup, so they can express breast milk and give it to the infant. You see, in that case, then the mother’s preference takes precedence, we just give it to the mother, the mother expresses and then gives it to the child. Yeah, so mother’s preference also is a determining factor apart from age.” (HCW 001)*

### Perceived benefits of pro/synbiotics and participation in the study

Four codes emerged that did not appear to be influenced by participant attributes such as age, education and employment status:

#### a) Accessible healthcare and education

This was the most coded node among the mothers/carers with 13 (92.8%) mentioning that they were reassured that both themselves and the participating infants would receive medical care when needed as it was stated as a benefit of study participation. Some of mothers/carers stated that education on good infant care practices during the trial visits had a positive influence:

> *“All I know is that since we got to the study, it has really helped me and made things easy for me because the child might be sick, and you are there to help out, so I have found everything so easy that even my husband feels the same way, so that is what I can say and what I have seen” (Mother 004)*

> *“Since she got into this study, it has really helped me, I can say it has really helped me a lot, because there was a time she could have fever, cold and there were also things that I dint know like how to take care of the baby’s umbilical cord, how to breastfeed, how to handle the baby and my own cleanliness, I dint know but you taught me … They also taught me how to bond with the baby well, how to dispose the baby’s diapers, and if in case you see danger signs when you should rush the baby to the hospital.” (Mother 001)*

#### b) Protection from diseases

Twelve mothers/carers (86%) perceived that the pro/synbiotics protected the infants from diseases:

> *“It prevents things like measles and worms that might be in her stomach” (Mother 003)*

> *“In my mind I believe that the supplement prevents some diseases that might affect the child” (Mother 004)*

> *“I have not seen my child contracting these normal diseases that children tend to have and her body is also good so those are the benefits that I can say” (Mother 007)*

> *“The difference that I am seeing between him and my other children is that the other ones were getting sick especially the age that this one is, but with him ever since I got to this study I have not seen him getting sick, he is healthy.” (Mother 008)*

#### c) Healthy growth of the infant

Nine mothers/carers (64%) stated that the pro/synbiotics helped their babies gain weight or made them stronger and more active:

> *“It makes the child strong, it keeps the baby healthy, the baby grows well” (Carer 002)*

> *“It helps the baby grow in the right way” (Mother 006)*

> *“when you compare it with other children, you find that they are less active and weak compared to this one who is active and strong” (Mother 013)*

#### d) Improved appetite

This node was coded during 7 (50%) of the mother/carer interviews:

> *“I can say it looks like something that provides the baby with vitamin and gives the child appetite making the child to breast feed well, so it doesn’t give you hard time, that is what I am seeing with the supplement” (Mother 004)*

> *“they started giving my child medicine and my child is breastfeeding really well” (Mother 008)*

### Perceived barriers to the intervention

Several codes emerged during analysis (fig 1). The commonly re-emerging codes were lack of paternal involvement, work and other commitments, other people’s opinions, challenges in delivery and lack of breastmilk. As with the perceived benefits, no correlations between participant attributes (e.g. age, education or employment status and number of children) and codes were observed.

**Figure 1:**
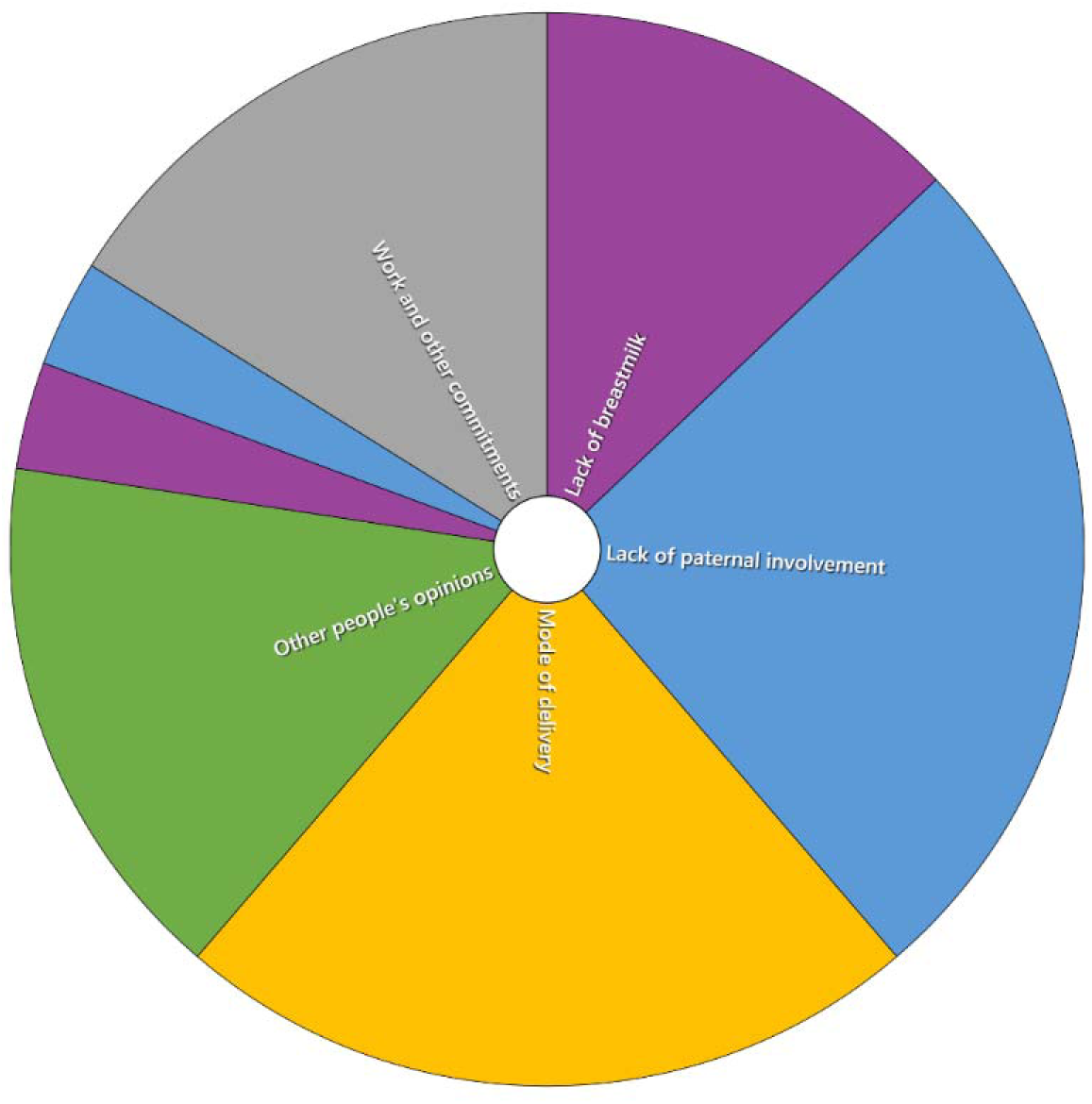
Distribution of codes under the perceived barriers theme compared by the number of items coded (mother/carer interviews)

### Lack of paternal involvement

Two mothers (17%) mentioned that fathers were absent during the consent process and did not have active involvement in taking care of the infants:

> *“[The father] is not saying anything because he doesn’t know that I am in this study so he just always sees people coming in the house” (Mother 012)*

In addition, two (29%) HCWs agreed that the lack of paternal involvement was a potential barrier to uptake of the pro/synbiotics. The following quote explains the issues that may arise because of this and solutions to overcome this barrier:

> *“The major challenge comes during introduction of the supplements to these participants is that; majority of the mothers who come to the facility don’t accompany themselves with their husbands. So, what you find is only a mother who has delivered in the ward. You will find the mother and the baby to consent but we tend to forget about the father. So it would be my advice that even if the father might be away, there is a role, a key role they play, so it would be prudent enough so they would be involved in this process, Because you find yourself, you introduce the study or the process to the mother, the mother agrees, but when they go home and they take supplements at one point, the father feels like he’s not part of the team. So that is one challenge - at any point we are introducing such an investigation of products, the two parties should be well informed. And if it’s a minor, the mother or the carer should be well informed about the procedures or activities that are ongoing.” (HCW 004)*

### Work and other commitments

Five (42%) mothers found it difficult to stick to the dietary administration and study schedules due to work and other personal commitments.

> *“I just think it’s just work, that is the thing that would make me find it difficult in following the rules of the study, I don’t see any other, because that is the only thing that would make me be away from the child” (Mother 004)*

In addition, 5 (42%) Peer Mothers also acknowledged commitments as an issue that would affect adherence to recommended breastfeeding practices. This was also a challenge when it came to carrying out their supportive roles as it would sometimes affect waiting times and inconvenience other household visits if the mother was not available at the scheduled visit.

> *“I have not seen any major challenge, is just that at times when we go for the visits you will find out that some people are not there, so you have to wait and at the same time you have to go and visit some other people that is where I find it challenging” (Peer mother 011)*

### Other community members’ opinions

Four (33%) mothers and 4 (33%) Peer Mothers raised concerns over the negative opinions of the neighbours and other community members. They stated that they were told their children were being exploited by the research staff who were conducting dangerous experiments on them.

> *“The thing I have found difficult is the opinion of others, relatives, friends, they say what will they benefit from you and what would you think of a person who takes full responsibility of your child, so they have not taken it as a good thing, yes they talk.” (Mother 013)*

> *“[A friend’s husband] was saying that the KEMRI only do the research with the child’s stomach and my baby will die.” (Mother 003)*

However, reassurance by the trial staff and coping strategies of the mothers/carers towards these negative comments seem to be good; some mothers said that they did not let these comments get to them and reply by saying that they are benefiting from the pro/synbiotics.

> *“If they come with the negative comments, I explain to them what I know about the study and how I was taught before I got to the study” (Mother 013)*

> *“With me, I am just okay because you cannot stop someone saying whatever they want to say, I just concluded that even if they want to talk let them talk, there is no problem” (Mother 014)*

There were concerns from both the mothers and Peer Mothers who stated that neighbours who witnessed trial staff entering the participants’ homes with cooler boxes perceived that the pro/synbiotics were anti-retroviral (ARV) medication for managing the Human Immunodeficiency Virus (HIV) and spread rumours that the infant and the mother were HIV positive. However, none of the key informants raised this point as a potential barrier.

> *“Initially, [the neighbours] thought that this food that we were giving were ARVs, so [the mother] would prefer you to hide it somewhere so that you can give the child because their neighbours would ask why the child is given medicine on a daily basis. So that is what we have tried to eradicate by telling them that it is food and not medicine, they were afraid at the beginning but now I see that they are okay” (Peer mother 008)*

### Challenges in the method of delivery

Challenges mainly related to the difficulties the mothers experienced with specific methods of administration, such as some content remaining in the container or the powder sticking to the infant’s throat when given directly.

> *“When given directly, at times [baby] finds it hard to consume it” (Mother 007) “When it is not mixed, there is some content of the medicine that remains in the bottle” (Mother 010)*

> *“When given directly, at times [the supplement] can stick on the throat and it is hard to digest.” (Mother 011)*

Four (33%) of the mothers stated that they were not able to express breastmilk either due to stigma or low milk production.

> *“There was another way we were told to use but I still dint have breast milk, we were told to use breast milk but since I did not have enough milk we were told to just use water.” (Mother 003)*

This was also raised by two Peer Mothers. In such instances, the mothers would be offered an alternative method of administration.

Key informants identified the HCW strikes and the COVID-19 pandemic as a challenge to data collection. These put strains on the health system in general as well as the trial:

> *“We’ve had several strikes, where the Ministry of Health staff was on strike. And that is a challenge to admission, and also the process that is required in the PROSYNK study. We also had stockouts, at times tests are required, but we are not able to do. We’ve been transferring the PROSYNK study’s patients, especially those who don’t require admission, to private hospitals and follow them up there for better care. And even in tests that are not able to be done within the hospital because of supply stockouts, we’ve always gone to private laboratories, and requested the test to be done. And that is a big challenge that we’ve experienced in the PROSYNK study.” (MOH 002)*

> *“So, the challenges that we’ve experienced so far is mainly with regard to conducting clinical trials in COVID-19 times. So that is top. Because right from when we started, there are instances where, like, at the moment, there is a surge, … we’ve divided the group into cohorts. A lot of planning and management has gone into this just to ensure that we always have a team in case there is exposure. This has also affected our recruitment, because when we are not working at full capacity, then we are not able to get participants. It could also complicate building follow ups. So, I think COVID-19 has really been something else for conduct of clinical trials.” (HCW 001)*

However, these challenges were considered to mainly affect study logistics rather than affecting community acceptability of the interventions and trial.

### Infant care practices

Understanding of recommended breastfeeding practices was generally good among mothers/carers. However, the understanding of the importance of breastfeeding practices seemed vague. Some mothers mentioned breastmilk protecting their babies from illnesses and helping them to grow stronger; however, most mothers said they practiced breastfeeding because breastmilk is the only type of food the baby can consume in early stages of life and they had relatively poor understanding of the benefits of breastmilk.

> *“The reason why the child is being breastfed is because that is the child’s food, just like you also, ugali is your food that even if you don’t take it you don’t feel comfortable, so I breastfeed the baby because that is his food that is on his mind.” (Mother 004)*

One carer mentioned the concept of connection with the baby while breastfeeding:

> *“Breast milk is stronger than that of the cow because that is just a cow but the mother has connection with the baby.” (Carer 005)*

Understanding of recommended WASH practices and importance of hygiene were well understood by the mothers/carers.

> *“Hygiene is important so that she cannot contract diseases which may make her have things like stomach ache” (Mother 006)*

> *“First for me, I have to ensure that his clothes are clean, his washed clothes are dry, secondly I as a mother who is breastfeeding, when I go somewhere and come back, If I have not bathe then I should clean my breast before breastfeeding, the environment should also be clean even if in any case someone wants to carry the baby, he/she should wash their hands or sanitize so that he/she doesn’t handle the baby with dirt” (Mother 004)*

Despite good understanding of WASH practices, Peer Mothers raised concerns over sometimes finding the infants in unsanitary conditions when they visit the households, especially when these visits were not expected.

> *“The places that we are visiting, it is not true that everybody is clean, some are clean and others are not. At times you visit and the mother has come out from the toilet and you find she will not remember to wash her hands, instead she will just go ahead and give [the supplement to the baby].” (Peer mother 003)*

One Peer Mother mentioned that this might be because of inaccessibility to the required facilities and products like soap in order to maintain good standards of hygiene.

> *“At times you find that when you go to some places you may find that it is difficult for the participant to even get soap, so to practice hygiene and cleanliness of her surroundings is hard so that maybe one of the causes” (Peer mother 006)*

## Discussion

Overall, there was a high level of satisfaction and acceptability regarding the use of the pro/synbiotics among all interviewed groups: mothers/carers, Peer Mothers and HCWs. Mothers/carers perceived healthy growth, prevention of diseases and improved appetite as benefits of the pro/synbiotics. This is consistent with previous findings of acceptability and perceived improved appetite and weight gain in studies of dietary tools or nutrients in western Kenya. In a qualitative study in rural and urban communities in western Kenya, dietary tools like slotted spoons, marked bowls and illustrative counselling cards to improve maternal and infant nutrition were well received by communities and mothers [24]. Similarly, high acceptability in focus group discussions was reported amongst Luo families in western Kenya, adults and children, in a study of a micronutrient powder (Sprinkles) [25]. Acceptability of lipid based nutrient supplements was also high in Haiti and Malawi although provided for older infants and children [26,27].

However, it is important to note that mothers/carers cited that the support they received as a consequence of participating in the clinical trial was the greatest benefit and Peer Mothers concurred in their recognition of this factor. Although there was high acceptance amongst mothers/carers of the study procedures, some found it difficult to stick to the weekly visit schedule due to work and other commitments. This may impair scaling-up interventions that require frequent administration especially amongst working mothers. Passing on the responsibility of pro/synbiotic administration to mothers/carers, after appropriate training and for those who feel confident, would reduce the number of scheduled visits and give mothers/carers more flexibility. However, compliance with pro/synbiotic administration would have to be monitored closely should pro/synbiotics be administered as a public health intervention.

The study also revealed varied views on the optimal method of pro/synbiotic administration. Most mothers/carers, Peer Mothers and HCWs agreed that it was easier to administer the pro/synbiotic directly into the mouth in older infants. For younger babies and some mothers, lack of breastmilk or difficulties expressing breastmilk to mix with the pro/synbiotic was an issue. Overall, HCWs reported that mothers/carers were able to administer the pro/synbiotics themselves. However, Peer Mothers raised concerns regarding the ability of mothers/carers to follow hygienic practice in pro/synbiotic administration.

A main barrier to pro/synbiotic administration was the negative influence of some community members. Extensive consultation with various stakeholders in Homa Bay county including the Community Advisory Board, religious leaders, local administrative officers, Peer Mothers, Community Health Volunteers and the Ministry of Health through the county and sub-county health management teams was undertaken before study start and also regularly throughout the study to answer any questions and respond to concerns. Our experience suggests that these concerted efforts to engage relevant stakeholders was critical to the success of the PROSYNK study. Similar stakeholder engagement would be required ahead of further research or use of pro/synbiotics as a public health intervention.

HCWs also highlighted the challenges posed by lack of paternal involvement. Although lack of paternal involvement was a recurring code under potential barriers, in families where both parents were involved in the care of the infant, mothers reported their partners’ involvement to be a positive and supportive factor. Engaging fathers in the consent for pro/synbiotic administration is likely important for both research and public health contexts.

Overall, the goal to reach saturation point was achieved under the various themes. On the other hand, despite the same codes re-emerging under the perceived barriers theme most of the time, focus group discussions or more key informant interviews may have identified new codes as this theme had the most variety of answers given during the interviews compared to the other themes.

### Limitations

We were careful when providing information for this qualitative research to explain that it was entirely separate from the main study. However, we cannot exclude the possibility that some mothers/carers may have felt intimidated or not fully at ease to share their honest feelings and opinions as they may have perceived the interviewer to be part of the trial team and any adverse comments may negatively affect their participation in the trial.

Additionally, having to translate the interview transcriptions from Swahili or Dholuo to English was a limitation. This meant that there was a possibility that some expressions that could not be directly translated might have been missed or misunderstood. The study was also conducted during the COVID-19 pandemic. Hence, interviews with the HCWs were done via WhatsApp calls. Following the interviews, the researcher noted that they found it harder to build rapport with the participants and that lack of visual cues may have hindered the potential of gaining more insights.

There was also a concern over equivalence as some key informant interviews were conducted by SZ researcher in English, whereas all interviews with mothers/carers and Peer Mothers were conducted in local languages by FA. A difference in positionality between the two interviewers might have led to misinterpretations while comparing the code trends between different participant groups (e.g. perceived benefits expressed by the mothers vs. key informants).

### Conclusions

Pro/synbiotic administration may be an affordable, safe and effective public health intervention to protect gut health and, thereby, reduce childhood malnutrition and its associated adverse outcomes. This study showed very high satisfaction and acceptability among community users (mothers/carers and Peer Mothers) as well as HCWs. It also revealed key issues regarding stakeholder engagement and processes for the administration of pro/synbiotics to infants by mothers/carers. Along with clinical and cost-effectiveness studies, further operational and qualitative research is required to explore ways to mitigate the barriers identified in this study.

## Acknowledgements

We thank the mothers/carers, HCWs and Peer Mothers for their valuable insights.

## Competing Interests

The authors declare that they have no competing interests.

## Data availability

All data produced in the present study are available upon reasonable request to the authors.

## Funding

The PROSYNK trial is funded by the Children’s Investment Fund Foundation; no additional funding was available for this qualitative research.

## Notes

### Competing Interest Statement

The authors have declared no competing interest.

### Clinical Trial

PACTR202003893276712

### Funding Statement

The PROSYNK trial is funded by the Childrens Investment Fund Foundation; no additional funding was available for this qualitative research.

### Author Declarations

Ethical approval was obtained from the Kenya Medical Research Institutes Scientific and Ethics Review Unit (KEMRI/SERU/CGHR/320/3917) and the Liverpool School of Tropical Medicine Research Ethics Committee (19-048).

